# Genome-wide Mendelian Randomization Identifies Potential Drug Targets for Dorsopathies

**DOI:** 10.1101/2024.05.01.24306675

**Authors:** Yu Cui, Jiarui Guo, Yuanxi Lu, Mengting Hu, He Zhou, Wancong Zhang, Shijie Tang

## Abstract

**Background:** Dorsopathies are a group of musculoskeletal disorders affecting the spinal column and related structures, contributing significantly to global disability rates and healthcare costs. Despite their prevalence, the genetic and biological mechanisms underlying dorsopathies are not fully understood.

**Method:** Summary-data-based Mendelian Randomization (SMR) and colocalization analysis were employed, using data from genome-wide association studies (GWAS) and cis-expression quantitative trait loci (cis-eQTLs) databases. Genes with a colocalization posterior probability (PP.H4) above 0.7 in SMR results were selected for additional analysis. These selected genes underwent MR analysis to examine possible causal connections with dorsopathies, and sensitivity analyses were carried out to ensure robustness. Additionally, two transcriptome-wide association studies (TWAS) were utilized to confirm and screen for potential drug targets.

**Result:** We identified four essential genes linked to dorsopathies: *NLRC4*, *CGREF1*, *KHK*, and *RNF212*. Mendelian randomization (MR) analysis revealed a potential causal link between these genes and dorsopathies. Elevated transcription levels of *NLRC4*, *CGREF1*, and *KHK* correlated with reduced dorsopathies risk, while increased levels of *RNF212* were associated with heightened risk of dorsopathies. Regarding methylation sites, an increase in *cg04686953* fully mediated the decreased risk of dorsopathies by *RNF212*. Similarly, the risk effect of *cg26638505* and *cg18948125* was entirely mediated by *NLRC4*, while *CGREF1* predominantly mediated the risk-increasing effect of *cg06112415* and the decrease effect of *cg22740783*.

**Conclusion:** Dorsopathies were associated with four pivotal genes: *NLRC4*, *CGREF1*, *KHK*, and *RNF212*. Methylation analysis identified cg04686953 and *cg22740783* as protective against dorsopathies risk, while *cg26638505*, *cg18948125*, and *cg06112415* exhibited a risk-increasing impact.

## 1 Introduction

Dorsopathies encompass a broad spectrum of musculoskeletal issues that impact the spinal column and its related structures[1]. These conditions are prevalent among individuals across various age groups and socioeconomic backgrounds, leading affected individuals to seek medical assistance. They significantly contribute to the burden of illness, disability, and distress. Since 1990, there has been a notable rise in the prevalence of these disorders, rendering them one of the primary causes of global disability-adjusted life years [2]. However, the pathogenic genes and biological mechanisms of the disease remain largely unknown. Research conducted on the general population has indicated a potential link between certain dorsopathies and lifestyle choices including smoking [3], a higher body mass index[4,5], and lack of physical activity [6]. Additionally, these dorsopathies have been found to coincide with various other health complications such as anxiety, depression, diabetes, cardiovascular, respiratory, and gastrointestinal ailments[7,8]. Furthermore, specific dorsopathies like osteopenia, osteomalacia, and tuberculosis can be influenced by factors such as diet, living environment, and psychological aspects. These particular disorders often occur alongside systemic comorbidities such as endocrine dysfunction and infection [8]. Relevant limitations in observational epidemiological studies include the potential for confounding, reverse causation, and diverse biases, which hinder comprehensive understanding of disease pathogenesis and identification of treatment targets.

However, the utilization of Mendelian randomization (MR) techniques, which rely on the random allocation of genetic variations, allows for the simulation of randomized controlled trials and can effectively mitigate the impact of confounding variables. In relation to research on dorsopathies, this implies a more precise evaluation of the causal association between drug targets and disease while excluding any factors that may potentially disrupt the findings [9]. MR utilizes genetic variants as instrumental variables to assess the causal impact of an exposure on outcomes. This approach has been extensively utilized in other research studies related to diseases and has effectively facilitated the identification of potential therapeutic targets for diverse medical conditions.[10] However, there is limited research on employing MR methods to explore potential drug targets for dorsopathies.

Therefore, to gain a deeper understanding of the pathogenesis of dorsopathies and identify more effective treatment approaches, further MR studies are needed to evaluate the drug targets for dorsopathies. This will help eliminate factors that could interfere with the results and offer new perspectives and methods for the treatment of dorsopathies. In theory, single nucleotide polymorphisms (SNPs) are distributed randomly and not impacted by environmental factors, making them an ideal tool for establishing causality. MR is a type of instrumental variable analysis that primarily employs SNPs as genetic instruments to determine the causal impact of an exposure (in this instance, circulatory proteins) on outcomes.[11]. Previous studies have successfully utilized MR to identify biomarkers and treatment targets for various diseases, such as aortic aneurysms [12], multiple sclerosis [13], and breast cancer [14].

However, in previous studies, the use of MR To analyze drug targets for dorsopathies is very rare. To address this research gap, our study focuses on utilizing genes as factors of exposure in drug target research, investigating the role of genes in dorsopathies development and their potential as viable drug targets. Genes have stable genetic characteristics, and they are unaffected by environmental factors, allowing for a more accurate assessment of the causal relationship between genes and dorsopathies [15]. By exploring dorsopathies’ potential drug targets through genes as the exposure factor, we aim to offer new perspectives and methods for the treatment and prevention of dorsopathies.

To investigate potential pathogenic genes related to dorsopathies, we employed a comprehensive analytical approach including summary-data-based Mendelian randomization (SMR) analysis, colocalization analysis, genome-wide association study (GWAS), and transcriptome-wide association study (TWAS). We utilized a meta-analysis dataset of cis-expression quantitative trait loci (cis-eQTLs) from peripheral blood samples as exposure data, along with results from the extensive FinnGen database. Following preliminary analysis, we identified candidate genes and established causal inference using MR methods. Additionally, we conducted mediation analysis of gene-mediated methylation sites to explore disease related methylation sites.

## 2 Method

### 2.1 Datasets

Summary-level data for the GWAS on dorsopathies were obtained from the FinnGen consortium, comprising 117,411 cases and 294,770 controls. This dataset represents the most recent GWAS findings and comprises the largest cohort of dorsopathies cases documented to date. The primary objective of FinnGen is to accumulate and rigorously analyze genomic and national health register data from 500,000 Finnish individuals.[16] In the context of drug development studies, we prioritized cis-eQTLs that were in closer proximity to the target gene. These cis-eQTLs used for SMR analyses were sourced from the eQTLGen Consortium [17] and the eQTL meta-analyses conducted on peripheral blood samples from a cohort of 31,684 individuals. For the TWAS analyses of eQTL data, we utilized the GTEx v8 European whole blood dataset. Given our specific emphasis on dorsopathies, we meticulously extracted comprehensive eQTL results exclusively from the whole blood samples within the GTEx dataset [18].

### 2.2 SMR analyses

We conducted SMR and heterogeneity in dependent instruments (HEIDI) tests analyses on cis regions using the SMR software (version 1.03) [19]. The methodologies for SMR analyses are detailed in the original work. In brief, SMR analyses employs a well-established MR approach. This technique employs a SNP at a prominent cis-eQTL as an instrumental variable (IV). The summary-level eQTL data serve as the exposure variable, and the GWAS data for a specific trait serve as the outcome variable. The primary objective is to explore a potential causal or pleiotropic association, wherein the same causal variant influences both gene expression and the trait. It is crucial to acknowledge that the SMR method lacks the ability to distinguish between a causal association, in which gene expression causally influences the trait, and a pleiotropic association, in which the same SNP affects both gene expression and the trait. This limitation arises because of the single instrumental variable (IV) in the MR method, which cannot differentiate between causality and pleiotropy. Nevertheless, the HEIDI test can make this distinction by discerning causality and pleiotropy from linkage. Linkage refers to cases in which two different SNPs in linkage disequilibrium (LD) independently influence gene expression and the trait. Although less biologically intriguing than causality and pleiotropy, the HEIDI test provides clarity in such scenarios. For the HEIDI test, a p-value below 0.05 was considered significant, suggesting that the observed association was attributable to linkage.

### 2.3 GWAS analyses

Multimarker analyses of genomic annotation (MAGMA) employs a multiple regression model to assess the cumulative effect of multiple SNPs within a specific gene region (±10 kb)[20]. The reference panel for calculating LD was derived from Phase 3 of the 1000 Genomes European population. Significance thresholds for GWAS analyses using both SMR and MAGMA were set at a false discovery rate (FDR) below 0.05, corrected using the Benjamini-Hochberg method.[21]

### 2.4 TWAS analyses

We conducted validations to integrate dorsopathies GWAS and eQTL data of whole blood from GTEx using the FUSION and UTMOST, widely utilized tools in prior TWAS investigations [22,23]. FUSION constructs predictive models using various penalized linear models, such as GBLUP, LASSO, Elastic Net, for the significant cis-heritability genes estimated from SNPs within 500 kb on either side of the gene boundary. Subsequently, it selects the optimal model based on the coefficient of determination (R2) calculated through a fivefold cross-validation. For UTMOST, we performed repeated 49 single-tissue association tests for each tissue. TWAS significance for both single-tissue analyses was determined with a Benjamini-Hochberg corrected FDR value below 0.05.

### 2.5 MR analyses

In conducting the two-sample MR analyses [24], we utilized the TwoSampleMR R package. The utilization of the two-sample MR framework requires employing two distinct datasets. In this study, genetic instruments, specifically cis-eQTL, served as exposures, while GWAS were utilized to determine outcome traits. The MR methodology investigates the relationship between gene expression and diseases or traits by employing genetic variants associated with gene expression as instrumental variables (exposure) and GWAS for the outcome measures. Mendelian Randomization facilitates exploration into whether alterations in gene expression causally impact diseases or traits. For instruments represented by a single SNP, we employed the Wald ratio. In cases where instruments consisted of multiple SNPs, we implemented the inverse-variance-weighted MR approach. When selecting SNPs, the significance thresholds were defined as P < 5 × 10^(−8) for genome-wide significance, with a linkage disequilibrium parameter (r^2) set to 0.1, and a genetic distance set to 10 MB.

### 2.6 Colocalization analyses

Conducted colocalization analyses using the coloc package in the R software environment (version 4.0.3). Colocalization analyses aims to assess the potential shared causality between SNPs associated with both gene expression and phenotype at a specific locus, thereby indicating the “colocalization” of these genetic signals. The analyses calculates posterior probabilities (PPs) for five hypotheses: H0 denotes no association with either gene expression or phenotype; H1 signifies an association solely with gene expression; H2 indicates an association exclusively with the phenotype; H3 suggests an association with both gene expression and phenotype through independent SNPs; and H4 implies an association with both gene expression and phenotype through shared causal SNPs. A substantial PP for H4 (PP.H4 above 0.70) strongly suggests the presence of shared causal variants influencing both gene expression and phenotype [25].

### 2.7 Methylation & Mediation analysis

We hypothesize that methylation sites exert influence on the pathogenic risk of dorsopathies through three primary pathways: a) they indirectly modulate the pathogenic risk by influencing gene expression; b) the methylation sites themselves have a direct impact on the pathogenic risk of dorsopathies; and c) methylation sites may affect the pathogenic risk of dorsopathies by influencing other confounders. It is crucial to underscore that the overall impact of methylation on the pathogenic risk of dorsopathies can be conceptualized as the combined action of these three pathways, expressed as T=a+b+c, where a denotes the indirect modulation of pathogenic risk by influencing gene expression, b denotes the direct effect of methylation sites on pathogenic risk, and c represents their influence on pathogenic risk through impacting confounding factors. Significance tests are conducted separately for T, x, y, and a. If all these parameters show significance, it can be inferred that the gene plays a role in the intermediate pathway, thereby substantiating the existence of the a) pathway.

## 3 Result

### 3.1 SMR analyses and colocalization for Preliminary Identifying Potential Genes

In our initial analyses phase, we utilized the eQTLGen dataset for SMR analyses to pinpoint genes significantly linked to dorsopathies. Employing the FDR method set our p-value significance threshold. The eQTLGen dataset provided extensive data, yielding 15,633 candidate genes.

We identified 269 genes significantly associated with dorsopathies (FDR_P < 0.05). Subsequent scrutiny uncovered four potential drug targets (*NLRC4*, *CGREF1*, *KHK*, *RNF212*) affecting dorsopathies through various intersecting methods. For clarity, Manhattan plots illustrated SMR results. Following gene identification, the HEIDI heterogeneity test (p_HEIDI > 0.05) sifted out genes lacking horizontal pleiotropy. SMR locus plots further elucidated gene outcomes.

We proceeded with colocalization analyses to merge GWAS and blood eQTL data for genes passing the SMR test. This aimed to determine if these genes colocalized with the dorsopathies trait. The colocalization test results robustly supported colocalization between the trait and all four genes (*NLRC4*: PP.H4 = 0.993, *CGREF1*: PP.H4 = 0.905, *KHK*: PP.H4 = 0.773, *RNF212*: PP.H4 = 0.923) meeting both SMR and HEIDI test criteria. Consequently, we identify these genes as top-priority candidates for subsequent functional studies.

### 3.2 MR analyses Validates Potential Gene Causal Relationships

Using cis-eQTL data from the eQTLGen Consortium, we conducted two-sample MR analyses on European summary statistics of individuals with dorsopathies. The discovery cohort, consisting of 117,411 cases and 294,770 controls from the FinnGen cohort, underwent inverse variance weighted (IVW) MR analyses to combine effect estimates from each genetic instrument. The analysis revealed associations between the genetically predicted expression of 956 genes and dorsopathies risk following multiple testing adjustments (FDR correction). Notably, *NLRC4* (OR = 0.886, 95%CI = 0.855-0.917, FDR_P = 1.09e-11), *CGREF1* (OR = 0.682, 95%CI = 0.585-0.794, FDR_P = 8.74e-07), *KHK* (OR = 0.936, 95%CI = 0.921-0.951, FDR_P = 3.67e-16), and *RNF212* (OR = 1.145, 95%CI = 1.056-1.241, FDR_P = 1.01e-03) were among the genes identified. To ensure the reliability of our findings, we conducted tests for horizontal pleiotropy, which did not reveal any evidence of its presence in the dataset. These additional analyses confirmed the absence of horizontal pleiotropy, bolstering the robustness and validity of our MR genetics findings.

### 3.3 Validation

#### 3.3.1 TWAS & UTMOST Validates Transcriptome-level Causal Relationships

In our quest to enhance causal inference and gain deeper insights into genetic associations with dorsopathies, we conducted a TWAS analyses on the four genes identified in previous analyses, utilizing Fusion and UTMOST software. This comprehensive approach aimed to elucidate the transcriptional associations of these genes with dorsopathies and bolster the evidence supporting their potential causal role.

The results revealed significant transcriptional associations for the four genes with dorsopathies, with all colocalization probabilities (PP.H4) exceeding 0.7. Specifically, elevated transcription levels of *NLRC4* (Z score = −5.33068, P = 9.78E-08), *CGREF1* (Z score = −4.7644, P = 1.89E-06), and *KHK* (Z score = −4.72939, P = 2.25E-06) were significantly correlated with a decreased risk of dorsopathies, whereas an increased transcription level of *RNF212* (Z score = 3.751506, P = 0.000176) was significantly associated with an increased risk of dorsopathies. Notably, these TWAS findings were consistent with those from the SMR analyses, providing robust support for the transcriptional associations of these genes with dorsopathies.

#### 3.3.2 MAGMA Validates Genome-level Causal Relationships

We also conducted a GWAS analyses using MAGMA software on the quartet of genes highlighted in preceding studies. This approach aimed to illuminate the associations of these genes with dorsopathies at the genome level, thereby reinforcing the evidence underpinning their potential causal involvement.

Based on the MAGMA analyses, we identified 855 significant genes that passed the FDR test. The outcomes unveiled noteworthy genome-level associations for the aforementioned genes with dorsopathies. Specifically, heightened transcription levels of *NLRC4* (FDR_P = 0.01851868), *CGREF1* (P = 0.002068503), and *KHK* (P = 0.009814817) exhibited significant correlations with reduced risk of dorsopathies, whereas increased transcription levels of *RNF212* (P = 0.00202954) were significantly linked with elevated risk of dorsopathies. Importantly, these GWAS findings corroborated those from the SMR analyses, thereby fortifying the robustness of the transcriptional associations of these genes with dorsopathies.

### 3.4 Methylation analysis & Mediation analysis

We evaluated the influence of *cg23387401* on *RNF212* expression (β = −0.52, P = 2.08e-14) and its link to dorsopathies risk (β = 0.19, P = 1.20E-07). From these findings, we identified how gene-mediated methylation impacts dorsopathies risk (β = −0.10, P = 1.35E-05), with an observed overall effect (β = −0.10, P = 2.68E-06) indicating a 93.79% intermediate effect proportion. This suggests that the rise in *cg23387401* is wholly mediated by *RNF212*, leading to reduced dorsopathies risk.

Likewise, we assessed *cg26638505*’s impact on *NLRC4* expression (β = −0.81, P = 1.25E-09) and its association with dorsopathies risk (β = −0.13, P = 7.87E-09). These calculations revealed the influence of gene-mediated methylation sites on dorsopathies risk (β = 0.11, P = 2.87E-05), with an overall effect (β = 0.09, P = 1.43E-03) and an intermediate effect proportion of approximately 120.00%. This suggests that cg26638505 elevation is entirely mediated by NLRC4, reducing dorsopathies risk.

We also analyzed *cg18948125*’s effect on *NLRC4* expression (β = −0.93, P = 1.72E-08) and its association with dorsopathies risk (β = −0.13, P = 7.87E-09). From this, we derived the impact of gene-mediated methylation sites on dorsopathies risk (β = 0.12, P = 5.50E-05), with an overall effect (β = 0.12, P = 5.59E-04) and an intermediate effect proportion of 104.41%. This suggests that *cg18948125* elevation primarily contributes to increased dorsopathies risk, mediated by *NLRC4*.

Similarly, *cg22740783’s* influence on *CGREF1* expression (β = 0.32, P = 7.71E-07) and its association with dorsopathies risk (β = −0.38, P = 2.12E-05) was calculated. This allowed us to identify the impact of gene-mediated methylation sites on dorsopathies risk (β = −0.12, P = 1.27E-03), with an overall effect (β = −0.13, P = 1.63E-04) and an intermediate effect proportion of 92.24%. Hence, we propose that the elevation in *cg22740783* is fully mediated by *CGREF1*, leading to decreased dorsopathies risk.

Lastly, we evaluated *cg06112415’s* impact on *CGREF1* expression (β = −0.34, P = 1.75E-06) and its association with dorsopathies risk (β = −0.38, P = 2.12E-05). Through these assessments, we identified the effect of gene-mediated methylation sites on dorsopathies risk (β = 0.13, P = 1.49E-03), with an overall effect (β = 0.12, P = 5.59E-04) and an intermediate effect proportion of 110.63%. This suggests that *cg06112415* elevation primarily contributes to increased dorsopathies risk, mediated by *CGREF1*.

## 4 Discussion

This study aimed to offer new perspectives and methods for the treatment and prevention of dorsopathies. In our research, the identification of candidate genes and the establishment of causal inference were conducted via MR methods, while the mediation analysis was used to explore disease related gene-mediated methylation sites. As a result, our study revealed four genes as potential therapeutic targets for dorsopathies, including *NLRC4*, *CGREF1*, *KHK* and *RNF212*. Among these genes, elevated transcription levels of the *NLRC4*, *CGREF1* and *KHK* were significantly associated with a decreased risk of dorsopathies, whereas a raised transcription level of *RNF212* was significantly related a higher risk of dorsopathies.

In order to find the novel drug targets for dorsopathies, we employed an integrative analysis that combined colocalization with MR to assess causal genes for dorsopathies. Later, we conducted tests for horizontal pleiotropy to ensure the reliability of our MR genetics findings. In addition, TWAS analyses on *NLRC4*, *CGREF1*, *KHK* and *RNF212* was conducted through Fusion and UTMOST software, which further illuminated the transcriptional associations of these genes with dorsopathies and bolstered the evidence supporting their potential causal role. Moreover, we conducted a GWAS analyses whose results were consistent with those from the SMR analyses, which reinforcing the evidence underpinning the potential causal association between these four genes with dorsopathies. Finally, we evaluated the influence of methylation sites on those genes expression and their links to dorsopathies. The results exhibited that the rise in *cg23387401* was entirely mediated by *RNF212*, the elevations of *cg26638505* and *cg18948125* were primarily mediated by *NLRC4*, and the up-regulation of *cg06112415* was fully mediated by *CGREF1*, leading to increased dorsopathies risk; while the rise of *cg22740783* was totally mediated by *CGREF1*, resulting in decreased dorsopathies risk.

NOD-like receptors (NLRs) family caspase activation and recruitment domain-containing protein 4 (*NLRC4*) gene encodes NLRC4, which is mainly expressed in macrophages, neutrophils, dendritic cells and glial cells[26]. As a member of the caspase recruitment domain-containing *NLR* family, NLRC4 partners with NLR family of apoptosis inhibitory proteins (NAIP) to assemble inflammasome complexes, which termed NAIP/NLRC4 inflammasomes[27]. Inflammasomes function as intracellular multi-protein platforms that are activated by pathogen-associated molecular patterns or damage-associated molecular patterns, triggering innate immune reactions and inflammatory caspase-activation to defense pathogens and danger signals and maintain the homeostasis[27–29]. The activation of caspases results in the proteolytic activation of the pro-inflammatory cytokines interleukin-1β (IL-1β) and/or interleukin-18 (IL-18)[27]. Meanwhile, activated caspase-1 also cleaves gasdermin D, whose N-terminal fragments become inserted into cell membranes, thereby facilitating the release of more IL-1β and IL-18[28,29]. In particular, inflammasomes were classified into canonical inflammasomes, which activating caspase-1, and non-canonical inflammasomes, which primarily activate caspase-4 and/or caspase-5 in human cells[29].

Specifically, the NAIP/NLRC4 inflammasome serves as a as part of the innate immune response and senses a range of intracellular bacteria and bacterial components in the host cell cytosol, such as *S. typhimurium*, *Legionella pneumophila*, *flagellin* and components of the virulence-associated type III secretion apparatus, thereby to mediate host defense against bacterial pathogens[26,27,30]. NAIP proteins function as specific cytosolic receptors for a variety of bacterial protein ligands[31], while NLRC4 interacts directly with caspase-1 to induce cell death, such as the pyroptosis of macrophage, and to cause rapidly initiate inflammation and vascular fluid loss[26]. The localized effects of NAIP/NLRC4 inflammasome could defend against bacterial pathogens. However, the aberrant activity of this cluster also leads to multiple clinical manifestations, including macrophage activation syndrome, neonatal enterocolitis and autoimmune disorders[28,31], and promotes the process of some diseases, such as gliomas[29], premature rupture of membranes[26], ulcerative colitis[28], rheumatoid arthritis[30] and so on. Sim et al. found that non-canonical pathway molecules of NAIP/NLRC4 inflammasomes, including caspase-4, caspase-5, and N-cleaved GSDMD, were significantly increased with each glioma grade[29]. Additionally, Zhu et al. reported that NLRC4 was upregulated in the membranes of patients with premature rupture of fetal membranes and recruited more caspase-1, which promotes the process of rupture of fetal membranes by inducing apoptosis and degrading extracellular matrix[26]. The study of An et al. presented that the persistent activation of NAIP/NLRC4 inflammasome induces macrophage pyroptosis mediated by caspase1-dependent cleavage of GSDMD and releases proinflammatory cytokines including IL-1b and IL-18, facilitating the occurrence and progression of UC[28]. In addition, research conducted by Delgado-Arévalo et al. indicated that NLRC4 as an inflammasome sensor differentially upregulated in CD1c+ cDC from patients with rheumatoid arthritis, and this sensor seems to be nonredundantly involved in the detection of intracellular dsDNA[30]. Nevertheless, our research observed the significantly inhibitory effect of up-regulated *NLRC4* expression on dorsopathies. Thus, further research on the roles and mechanisms of *NLRC4* and inflammatsome in the progress of dorsopathies is needed.

Cell growth regulator with EF-hand domain 1 (*CGREF1*) gene encodes a novel secretory protein with 2 Ca2+-binding EF-hand domains which plays an important role in eukaryotic cellular signaling[32]. *CGREF1* mRNA has high expressions in HCT116, H1299 and HepG2 cells while expressing at low levels in other cell lines including Raji, Jurkat, BT325, PC12[32]. Mechanistically, *CGREF1* is regulated by p53[33], and the overexpression of *CGREF1* significantly inhibits the transcriptional activity of AP-1, reduces the phosphorylation of ERK (extracellular signal-regulated kinases) and p38 MAPK (mitogen-activated protein kinases), and suppresses the proliferation of HEK293T and HCT116 cells[32]. Furthermore, *CGREF1* can decrease the percent of G2/M and S phase and repress cell proliferation while overexpressing[32]. We found that the increased expression of *CGREF1* is significantly related to the decreased risk of dorsopathies. In the research of Xiang et al., the colorectal cancer patients with higher expression of *CGREF1* were found to have significantly better overall survival than patients with lower expression, and the role of *CGREF1* in the prognosis of Early-onset colorectal cancer was reported[33]. Furthermore, Xie et al. observed that *CGREF1* expression were high in the tissue of osteosarcoma patients while it were lowly expressed in normal tissue, and speculated that *CGREF1* can predict drug resistance to osteosarcoma[34]. However, the biological function of *CGREF1* is poorly explored, and further study for its mechanisms and roles in these diseases is warranted.

Ketohexokinase (*KHK*) is an enzyme that phosphorylates fructose to produce fructose-1-phosphate (F-1-P) at the first rate-limiting step in fructose metabolism[35,36]. The gene *KHK* encodes two isoforms, KHK-C and KHK-A. Considered as the primary enzyme in fructose metabolism, KHK-C is exclusively expressed in a few tissues, especially in the liver, and drives the aforementioned reaction rapidly, resulting in the accumulation of uric acid and transient depletion of intracellular phosphate and ATP[35–38]. In the contrary, KHK-A has a much weaker affinity and a higher Michaelis constant for fructose[38]. However, KHK-A can directly phosphorylate phosphoribosyl pyrophosphate synthetase 1 (PRPS1) by prevention of inhibitory nucleotide binding and facilitation of ATP binding[37]. As a result, KHK-A improves nucleic acid synthesis and promotes the G1/S phase transition in the cell cycle, accelerating cell proliferation[36,38].

Recent studies have highlighted the importance of *KHK* as the key mechanism stimulating the various adverse metabolic effects of fructose, such as impaired insulin sensitivity, hypertriglyceridemia, and oxidative stress[35]. In addition, fructose metabolism seems to provide cancer cells with the supplementary fuel required for proliferation and metastasis in colon cancer, and glioma[36,38]. Moreover, alternated expression of *KHK* is also related to several diseases. For instance, knocked down the expression of *KHK* dramatically reduced fructose-induced production of reactive oxidative species (ROS) in proximal tubular cells[35], indicating the association between KHK and ROS. Furthermore, KHK-A activates and upregulates PRPS1, which could lead to enhanced nucleic acid synthesis for tumourigenesis[38]. Kim et al. reported that most cancer cell lines predominantly expressed KHK-A rather than KHK-C, and KHK-A overexpression could augment cell invasion[36]. Moreover, in their study, upon fructose stimulation, KHK-A acted as a nuclear protein kinase and triggered Epithelial-mesenchymal transition in breast cancer, promoting breast cancer metastasis[36]. Besides, loss-of-function variants in *KHK* can cause essential fructosuria, an autosomal recessive disease characterized by intermittent appearance of fructose in the urine[39]. Yang et al. found the the overexpression of KHK-A in cell lines of oesophageal squamous cell carcinoma, which may finally promote the proliferation, tumourigenicity and motility of ESCC cells[38]. Similarly, KHK-A was considered to play an instrumental role in promoting de novo nucleic acid synthesis and hepatocellular carcinoma development[37]. In our study, the raised expression of *KHK* is found to be positively related to decreased risk of dorsopathies. In oder to understand the mechanism of changed expression level of *KHK* in dorsopathies, further research are needed.

Regulatory factor X2 (*RFX2*) gene is essential for maintaining normal spermatogenesis and involved in spermatogenesis impairment and male infertility in mice[40]. In particular, Ring finger protein 212 (*RNF212*) gene is important for crossing over and chiasma formation during meiosis[41]. Mouse *Rnf212* has a central role in designating crossover sites and coupling chromosome synapsis to the formation of crossover-specific recombination complexes. In humans, *RNF212* has been associated with variation in the genome-wide recombination rate[41]. The protein encoded by *RNF212* has homology to two meiotic procrossover factors: Zip3 and ZHP-3[42]. It functions to couple chromosome synapsis to the formation of crossover-specific recombination complexes[43]. Moreover, with the symbolic RING-finger domains, *RNF212* protein is a RING-family E3-ligase for Small ubiquitin-like protein (SUMO), and the latter plays an important role in assembly and disassembly of synaptonemal complex by regulating protein–protein inter action during meiosis[42,43,43]. In addition, *RNF212* stabilizes association of a subset of MutSγ complexes with recombination sites[42,44]. MutSγ complex is a kind of meiosis-specific recombination factors, working as an attractive target for non-crossover/crossover differentiation[42]. It binds and stabilizes DNA strand-exchange intermediates to promote both homolog synapsis and crossingover[44]. In mammals, every pair of chromosomes obtains at least one crossover, while the majority of recombination sites yield non-crossovers. Non-crossovers are inferred to arise from the disassembly of D-loops and annealing of DNA double-strand breaks ends in a process termed synthesis dependent strand annealing[42]. Designation of crossovers involves the formation of metastable joint molecules and selective localization of SUMO-ligase *RNF212* to a minority of recombination sites where it stabilizes pertinent factors, such as MutSγ[42,44]. Furthermore, this differential *RNF212*-dependent stabilization of key recombination proteins at precrossover sites is thought to be the basic feature of crossover/non-crossover differentiation[42]. It is suggested that *RNF212*-mediated SUMOylation may stabilizes the association of MutSγ with nascent crossover CO intermediates in a number of ways, such as promoting protein-protein interactions, altering ATP binding and hydrolysis (which modulate the binding and dissociation of MutSγ complexes) or antagonizing ubiquitin-dependent protein turnover[41,42].

Insufficient RNF212 accumulating at recombination sites can lead to crossing-over stochastically fails[43]. Fujiwara et al. reported that *Rnf212* knock out (KO) in mice leads to infertility of male and female due to the loss of SPCs(spermatocytes) at post-anaphase stage. Moreover, crossing over is diminished by ≥90% in *Rnf212*−/− mice[43]. In particular, the *Rnf212* KO spermatocytes lack chiasmata and exhibit depletion of spermatids and mature spermatozoa[41]. A nonsense mutation in the *Rnf212* gene was discovered in repro57 mutant mice, and this mutant mice exhibited male infertility, arrest of spermatogenesis in meiosis, and defects in cytological markers of recombination and chiasma formation, which is similar to the *Rnf212* KO phenotype[41]. In humans, the rate of crossing-over varies significantly between individuals, and higher maternal crossover rates have been associated with greater fecundity[42]. However, in the absence of RNF212, designation of crossover sites fails because no MutSγ complexes are stabilized beyond early pachynema[44]. Yu et al. detected that the frequencies of allele C and the genotype CC at the *rs4045481* locus in *RNF212* gene were significantly higher in patients with azoospermia in comparison with controls. Furthermore, they reported that homozygous of allele C (genotype CC) may decrease the activity of pre-mRNA due to the disappearance of the binding motifs of SRSF5, leading to the reduced expression of *RNF212* and influencing normal spermatogenesis, consequently increasing ther risk of azoospermia[40]. Nevertheless, we found that the increased expression of *RNF212* exhibited a positive correlation with dorsopathies. The influence of raised expression of *RNF212* is poor classified, and further researches are required to illustrate the mechanism of increased *RNF212* level and dorsopathies.

To summary, our study revealed the causal associations between the genetically predicted expression of four genes (*NLRC4*, *CGREF1*, *KHK* and *RNF212*) and dorsopathies risk, which offers new perspectives and strategies for the improved diagnosis, treatment and prevention of dorsopathies.

Based on our results, we hypothesized that increased expression level of *NLRC4*, *CGREF1* and *KHK* are associated with decreased risk of dorsopathies, while the increased expression level of *RNF212* has the opposite effect. Moreover, further study focusing on the mechanism of these expression changes in dorsopathies are needed, and future clinical studies should be conducted.

## Data Availability

The original contributions presented in the study are included in the article/Supplementary material, further inquiries can be directed to the corresponding authors.

## 5 Acknowledgements

The authors appreciate the publicly available data of the FinnGen consortium, the eQTLGen consortium, the GTEx project, and the MRC IEU OpenGWAS database.

## 6 Conflict of interest

The authors declare that the research was conducted in the absence of any commercial or financial relationships that could be construed as a potential conflict of interest.

## 7 Funding

The study and publishing of this article were not supported by any funding.

## 9 Tables

**Table 1.** TWAS / SMR/HEIDI results of the GWAS data on Dorsopathies, blood eQTL data.TWAS/SMR/HEIDI results of the GWAS data on Dorsopathies, blood eQTL data. Ch represents chromosome;position indicates the gene’s position, Z_TWAS, P_TWAS are the Z-score and p-value of the TWAS test; Tissue represents the tissue source used in the TWAS analyses. topSNP represents the top SNP in the SMR analyses. P_SMR is the p-value for the SMR test; B_SMR is the effect size from the SMR test; SE_SMR is the standard error of B_SMR; P_HEIDI is the p-value for the HEIDI test; N_HEIDI is the number of SNPs used in the HEIDI test; FDR represents false discovery rate.

| Gene | Ch | Position | Z_TWAS | P_TWAS | P_TWAS (FDR) | Tissue | topSNP | B_SMR | SE_SMR | P_SMR | P_SMR (FDR) | P_HEIDI | N_HEIDI |
| --- | --- | --- | --- | --- | --- | --- | --- | --- | --- | --- | --- | --- | --- |
| NLRC4 | 2 | 32265853 | -5.33 | 9.8E-08 | 5.1E-05 | GTEXv8.Whole_Blood | rs385076 | -0.13 | 0.02 | 7.9E-09 | 9.5E-06 | 3.6E-01 | 20 |
| CGREF1 | 2 | 27119114 | -4.76 | 1.9E-06 | 5.0E-04 | GTEXv8.Whole_Blood | rs2304681 | -0.38 | 0.09 | 2.1E-05 | 4.6E-03 | 4.4E-01 | 20 |
| KHK | 2 | 27086746 | -4.73 | 2.3E-06 | 5.7E-04 | GTEXv8.Whole_Blood | rs2119026 | -0.06 | 0.01 | 2.3E-06 | 1.0E-03 | 2.5E-01 | 20 |
| RNF212 | 4 | 1113561 | 3.75 | 1.8E-04 | 0.01 | GTEXv8.Whole_Blood | rs4974591 | 0.19 | 0.04 | 1.2E-07 | 9.4E-05 | 5.5E-02 | 20 |

**Table 2.** Main Results of Intermediary analyses. b is the effect size, se is the standard error of effect size, and p is the P-value. prop_mediated represents the proportion of indirect effect in the total effect.

| Marker | Gene | Methylation<br>on<br>Gene |  |  | Methylation<br>on<br>Dorsopathies |  |  | Gene<br>on<br>Dorsopathies |  |  | Intermediary analyses |  |  | prop<br>mediate<br>d | prop<br>mediated<br>se |
| --- | --- | --- | --- | --- | --- | --- | --- | --- | --- | --- | --- | --- | --- | --- | --- |
|  |  | b | se | p | b | se | p | b | se | p | indirect | indirect_se | indirect_p |  |  |
| cg0468695<br>3 | RNF21<br>2 | -<br>0.5<br>2 | 0.0<br>7 | 2.08E<br>-14 | -0.1 | 0.0<br>2 | 2.68E<br>-06 | 0.1<br>9 | 0.0<br>4 | 1.20E<br>-07 | -0.1 | 0.02 | 1.35E-05 | 93.79% | 0.29 |
| cg2663850<br>5 | NLRC4 | -<br>0.8<br>1 | 0.1<br>3 | 1.25E<br>-09 | 0.0<br>9 | 0.0<br>3 | 1.43E<br>-03 | -<br>0.1<br>3 | 0.0<br>2 | 7.87E<br>-09 | 0.11 | 0.03 | 2.87E-05 | 120.00<br>% | 0.47 |
| cg1894812<br>5 | NLRC4 | -<br>0.9<br>3 | 0.1<br>7 | 1.72E<br>-08 | 0.1<br>2 | 0.0<br>3 | 5.59E<br>-04 | -<br>0.1<br>3 | 0.0<br>2 | 7.87E<br>-09 | 0.12 | 0.03 | 5.50E-05 | 104.41<br>% | 0.4 |
| cg2274078<br>3 | CGRE<br>F1 | 0.3<br>2 | 0.0<br>6 | 7.71E<br>-07 | -<br>0.1<br>3 | 0.0<br>4 | 1.63E<br>-04 | -<br>0.3<br>8 | 0.0<br>9 | 2.12E<br>-05 | -0.12 | 0.04 | 1.27E-03 | 92.24% | 0.38 |
| cg0611241<br>5 | CGRE<br>F1 | -<br>0.3<br>4 | 0.0<br>7 | 1.75E<br>-06 | 0.1<br>2 | 0.0<br>3 | 5.59E<br>-04 | -<br>0.3<br>8 | 0.0<br>9 | 2.12E<br>-05 | 0.13 | 0.04 | 1.49E-03 | 110.63<br>% | 0.47 |

## 10 Figures

**Fig. 1.**
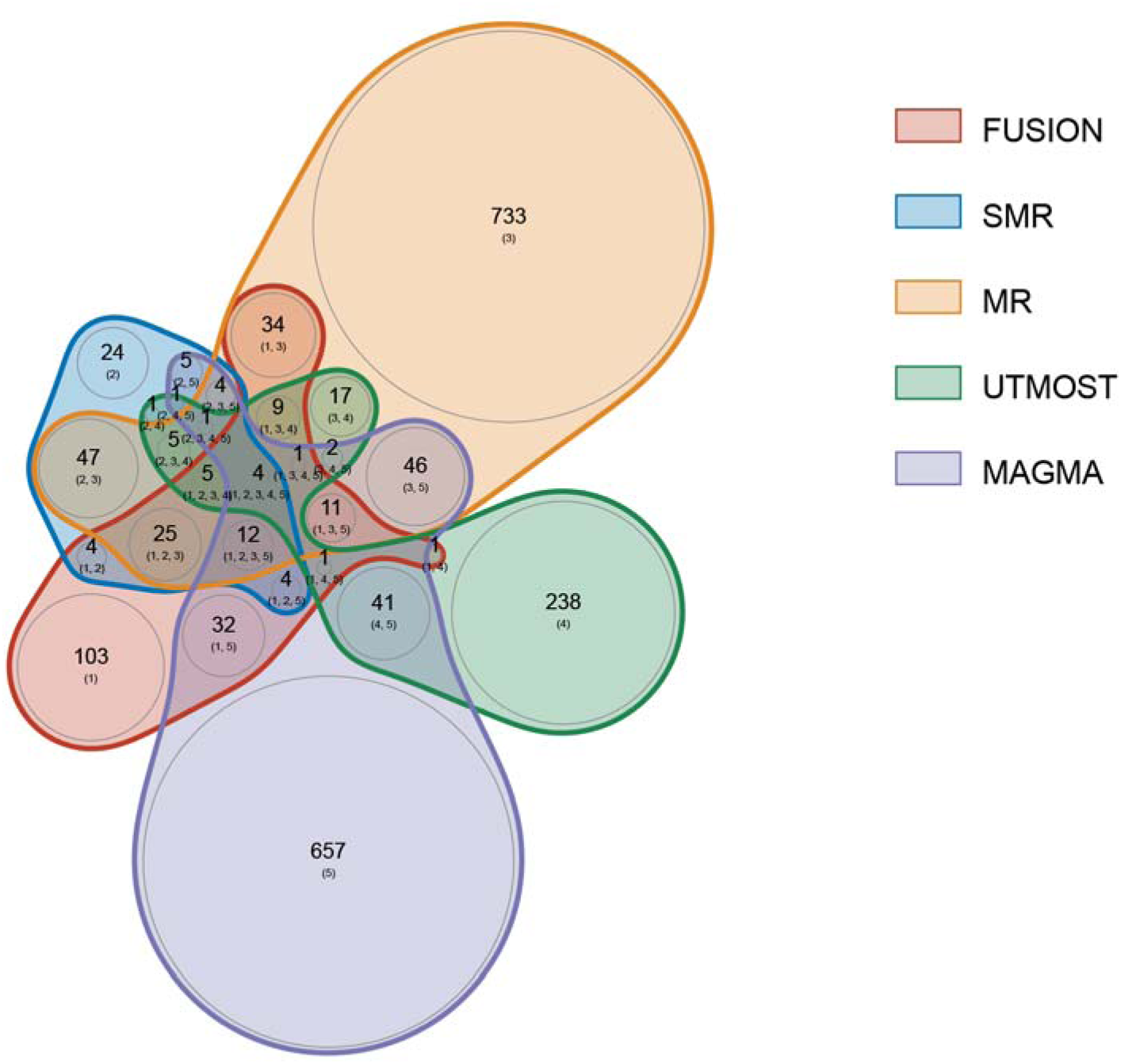
Venn Plot of the main results of multi analysis

**Fig. 2.**
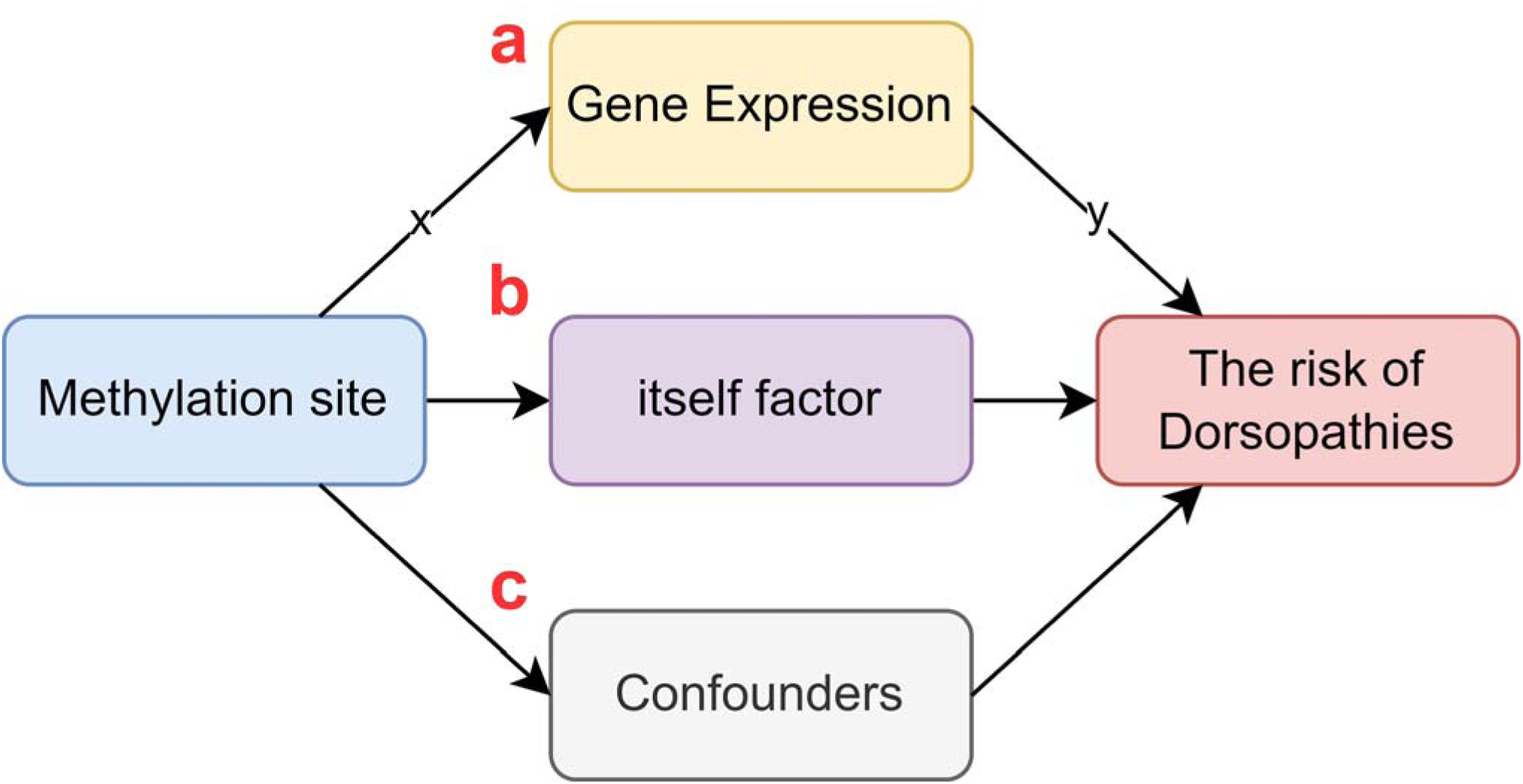
Methylation sites affect disease pathways.

**Fig. 3.**
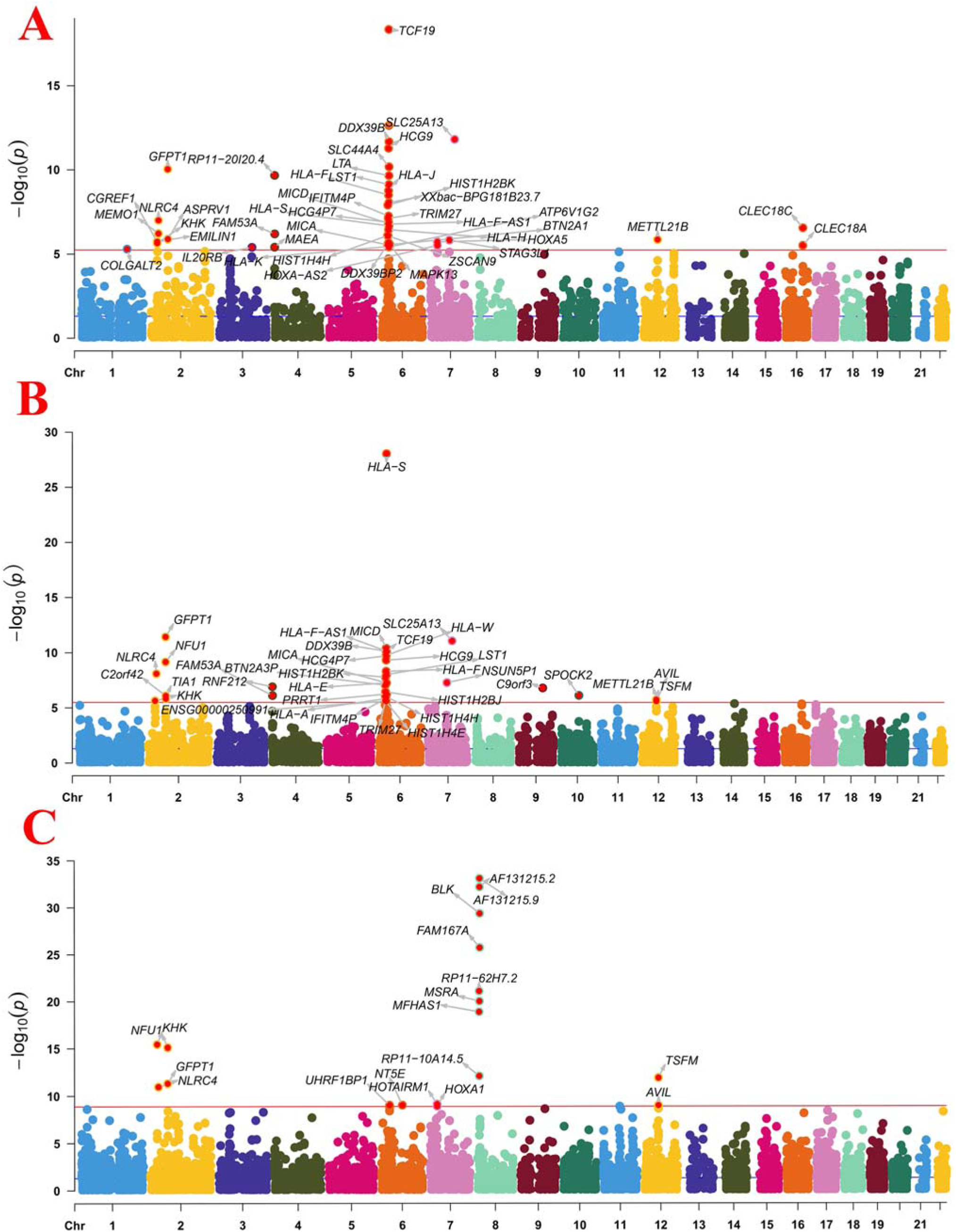
Manhattan plot of the MR/SMR/FUSION analysis results using QTLs and Dorsopathies GWAS summary statistics. The red dashed line represents the Bonferroni-corrected significance threshold.

**Fig. 4.**
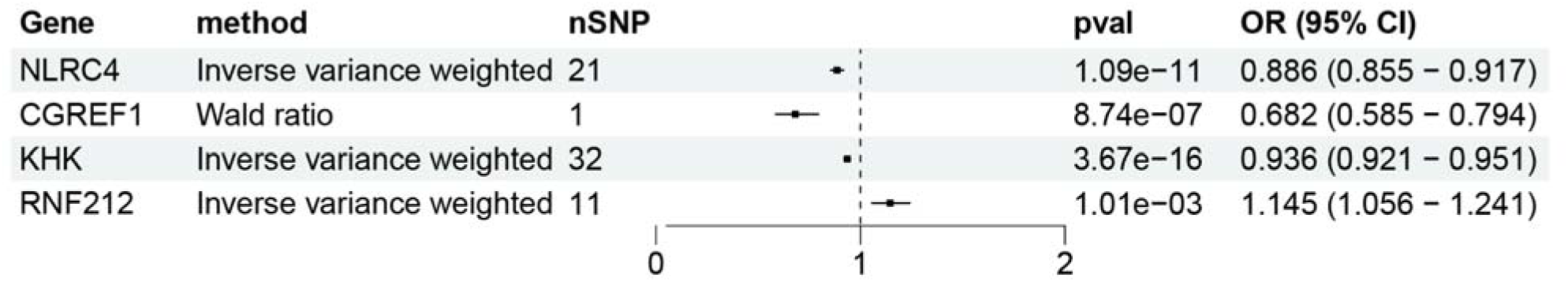
MR results for genes expression significantly associated with dorsopathies after FDR correction.

**Fig. 5.**
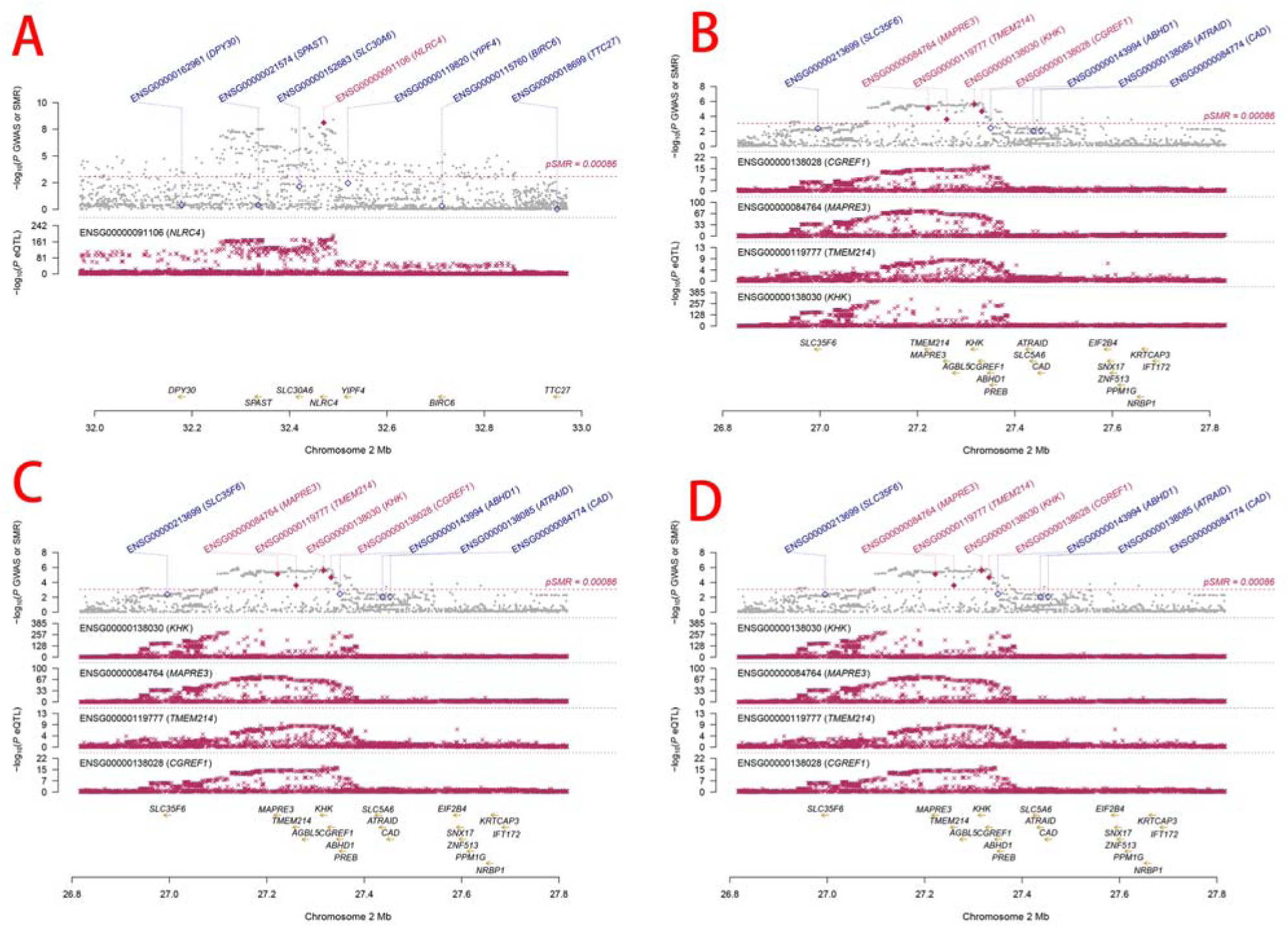
SMR locus plot illustrating dorsopathies at the gene locus utilizing blood eQTL data. In the upper plot, each gray dot represents a SNP identified through GWAS on dorsopathies. A red diamond indicates passage of the SMR test for the probe, while a solid diamond signifies successful completion of both the SMR and HEIDI tests. In the lower plot, each red cross represents an SNP identified in the eQTL study corresponding to each gene. The x-axis displays the genomic positions (Mb, GRCh37) of SNPs, probes, and genes on the chromosome. The y-axis displays the negative logarithm (base 10) of p-values for SNPs identified in the GWAS on dorsopathies, SMR test, and eQTL study corresponding to each gene.

**Fig. 6.**
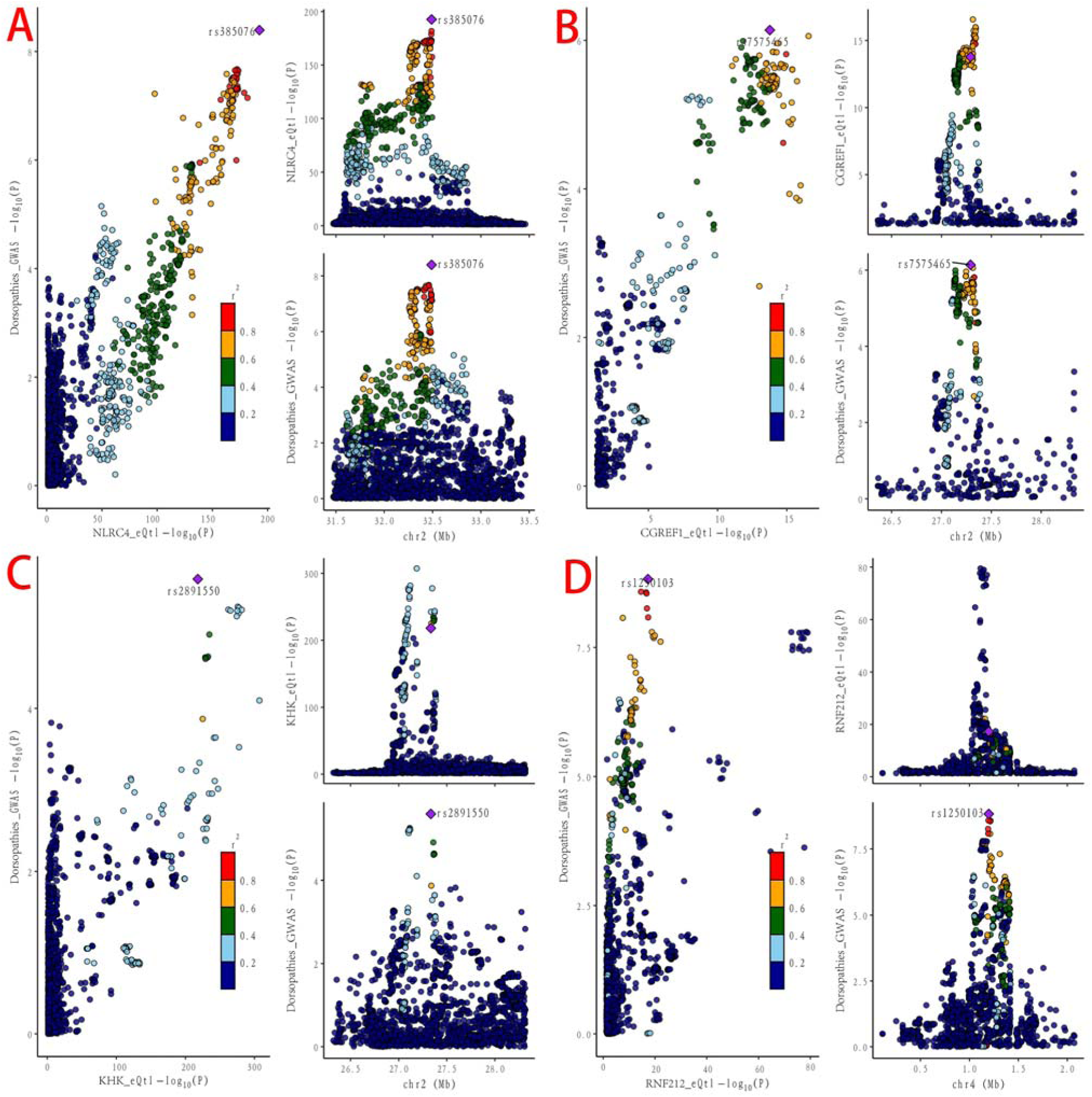
Locus comparison plot displays the results of colocalization analysis for SNPs associated with gene expression in blood and datasets related to dorsopathies. Each dot represents a specific SNP, with the color indicating its linkage disequilibrium (LD) value (r2) with the lead GWAS variant, marked by a purple diamond. In the right panel, the x-axis represents genomic positions in megabases (GRCh37) along the chromosome, while the y-axis shows the -log10 p-values for SNPs from the dorsopathies GWAS (top) and the gene expression eQTL study (bottom). The left panel compares the p-values from the dorsopathies GWAS with those from the gene expression eQTL study.

## Notes

### Competing Interest Statement

The authors have declared no competing interest.

### Funding Statement

This study did not receive any funding

### Summary of Updates

author affiliations updated, revised some affiliations for the first 5 authors。

